# The Effects of AI-Guided Exercise and a Smart Ring on Arterial Stiffness (GONDOR-AS): protocol for a randomized controlled trial

**DOI:** 10.64898/2026.03.19.26348812

**Authors:** Heikki Pentikäinen, Sofia Strömmer, Drake Caraker, Julia Kosonen, Aleksi Rantanen, Saila Hiltunen, Pirjo Komulainen, Heidi Similä, Massimiliano de Zambotti, Kai Savonen, Pauli Ohukainen

## Abstract

**Background:** Cardiovascular disease (CVD) prevention is limited by the major challenge of low long-term adherence to effective lifestyle regimens. Arterial stiffness (measured by carotid-femoral pulse wave velocity, cfPWV) and maximal cardiorespiratory fitness (measured by VO_2_max), are modifiable risk factors for CVD but require sustained lifestyle change. Wearable technology provides continuous measurement and offers a scalable platform to deliver health interventions. A combination of continuous monitoring with a wearable device and an artificial intelligence (AI) -based coach personalized for individual data and preferences could be a powerful, low-barrier tool for achieving sustainable cardiovascular health benefits by directly addressing the adherence challenge.

**Objective:** We will study the comparative effectiveness of a wearable and an interactive app-based AI coaching intervention promoting moderate exercise on improving gold-standard cfPWV and VO_2_max. This will be compared to a supervised high-intensity interval training (HIIT) group (benchmark with known benefits for VO_2_max) and a control group using only Oura Ring (passive monitoring). We will also conduct a detailed Process Evaluation (structured interviews) to study the feasibility and experience of interacting with the AI coach.

**Methods:** This randomized controlled trial recruited 165 eligible sedentary participants aged 30-65 years. Co-primary outcomes cfPWV and VO_2_max were measured at baseline and will be repeated after 12 weeks. Participants were equally randomized into three groups: an AI-based coaching group (steady-state exercise), a HIIT group (supervised exercise) and a control group (usual low activity). The AI-based coaching group receives personalized guidance using large language model (LLM) technology. All participants wear Oura Ring and are blinded to cardiovascular health metrics provided by the ring.

**Results:** The recruitment for the study began in October 2024 and will end when 165 men and women have been recruited. Data collection for the study is scheduled to conclude early 2026. Data collection is ongoing.

**Conclusions:** This study will evaluate if a highly scalable, AI-based coaching intervention can achieve comparable gains in CV structural health (cfPWV) and functional capacity VO_2_max relative to a resource-intensive supervised HIIT benchmark. The findings will provide essential evidence on the use of digital health tools to promote sustainable exercise adherence.

**ClinicalTrials.gov registration identifier:** NCT06644014 (Registered: 2024-10-15)

## Introduction

Cardiovascular disease (CVD) is a leading cause of death, disability, and morbidity worldwide [1]. While chronological age is a major determinant of CVD, the key modifiable factors remain lifestyle-related. Two significant, lifestyle-related, modifiable risk factors are cardiorespiratory fitness (measured by maximal oxygen consumption; VO_2_max) [2] and arterial stiffness (measured by carotid-femoral pulse wave velocity, cfPWV) [3]. Cardiovascular age (CVA) is an emerging metric which expresses an individual’s CV risk in units of chronological age [4]. It provides an intuitive and actionable measure of health and is heavily influenced by cfPWV. It is also negatively correlated with VO_2_max [5]. Aerobic exercise consistently reduces arterial stiffness [6] and improves VO_2_max but long-term adherence to exercise regimens remains a major challenge for many individuals.

Wearable technology is an emerging and scalable solution to enable continuous monitoring of one’s health and possibly to overcome the challenge of adopting new health habits, including regular exercise [7]. Several randomized trials have shown that interacting with an LLM-based chatbot is efficacious for increasing physical activity [8]. However, these studies rely on standalone discussions without real-time context from wearable-derived physiological signals and the ensuing feedback-loop for active engagement. Wearable devices like Oura Ring utilize multiple sensors to provide continuous, at-home estimates of e.g. physical activity, CVA and sleep quality. The rise of artificial intelligence (AI) enables moving from measurement to using the device as a platform for interventional delivery [9]. AI-based coaching may offer continuous, low-barrier, personalized health behavior guidance that is adaptable to the user’s measured data, life circumstances, and preferences, directly addressing the adherence challenge.

In this trial, the AI-based coaching system functions as a consumer-facing digital health intervention aimed at supporting individuals in adopting and maintaining physical activity behaviors known to reduce arterial stiffness. It delivers personalized guidance, motivation, and feedback based on physiological and behavioral data collected by Oura Ring, together with limited self-reported information. The system is intended for use by adults from the general population seeking to improve cardiovascular health outside formal healthcare settings. Within the trial, it represents one of the behavior-change strategies being compared to evaluate its efficacy in reducing arterial stiffness. The AI system does not inform clinical decision-making or patient management. Instead, it provides automated, participant-facing support for lifestyle modification.

The purpose of this randomized controlled trial (RCT) is to evaluate the effectiveness of an AI-based coaching intervention promoting moderate, steady-state exercise on improving both gold-standard cfPWV and VO_2_max. This takes place via a feedback-loop where the user can freely interact with the AI and gain real-time insights drawn from their data. The study employs a three-arm parallel design with two comparisons. The supervised high-intensity interval training (HIIT) group serves as the benchmark - a comparison whose well-documented advantage is a relatively rapid gain in VO_2_max. This group tests what is physiologically possible but might be challenging to adhere long-term, especially in the broad general population. The Oura Ring Only group serves as the baseline comparator to isolate the effect of the AI coaching beyond the benefits of passive self-monitoring via the wearable. Primary outcomes are the change in cfPWV and VO_2_max at 12 weeks. Process Evaluation (interviews and chat logs) is conducted for understanding the qualitative aspects of feasibility and mechanism of adherence for the novel AI coach.

## Methods

### Study design

Guidance with Oura and AI for Reducing Arterial Stiffness (GONDOR-AS) is a 12-week RCT (ClinicalTrials.gov identifier: NCT06644014) consisting of three groups (N = 55 per group):

- Oura Ring only (a control group)
- Oura Ring + HIIT twice a week
- Oura Ring + AI-based coaching for steady-state aerobic training (no external supervision)

All groups will be blinded to their CVA/PWV information but will use identical Oura Ring devices. Apart from the AI coach group, Oura App experiences will also be identical.

The study will be implemented in collaboration with Oura company and Kuopio Research Institute of Exercise Medicine (KuLTu). Patients and the public were not involved in the initial design of this academic-industry collaboration protocol.

### Recruitment of the study participants

The data collection for this study was initiated immediately after receiving approval from the Regional Medical Research Ethics Committee of Eastern Finland Collaborative Area. We used internal recruitment channels of Kuopio University Hospital, University of Eastern Finland and Savonia University of Applied Sciences to recruit personnel and students of these institutions. Kuopio city recruitment channels, newspapers and social media (eg. LinkedIn and Facebook) were also utilized to recruit participants from the general population. Altogether, 165 participants, aged 30-65 years, will be recruited to the study.

### Inclusion and exclusion criteria

Inclusion criteria were 30-65 years of age and self-reported moderate intensity physical activity less than 2 hours 30 minutes per week or vigorous intensity physical activity less than 1 hour 15 minutes per week. This agrees with international physical activity guidelines [10] for the minimum amount of recommended aerobic physical activity per week for adults. Self-reported physical activity data were subsequently reviewed by an expert researcher (HP), who verified and finalized participants’ activity profile classifications based on the reported information. Exclusion criteria were coronary artery disease, diabetes, uncontrolled hypertension or another condition which could compromise the safe participation to the exercise intervention evaluated by a physician (KS). Inability to use Oura Ring (e.g. work-related restrictions) was also considered an exclusion criteria; however, short interruptions (e.g. 1-2 hour work tasks without ring) were allowed.

If a participant experiences an adverse event or injury that compromises safe participation, their assigned intervention will be discontinued immediately upon review by the study physician (KS). They will remain in the study for final follow-up and be analyzed under the Intention-to-Treat principle (see below).

### Study measurements

Study assessments will be arranged in two measurement visits both at baseline and after 12 weeks intervention at KuLTu. Oura Ring will be provided at the first measurement visit approximately two weeks before the start of the intervention. This is necessary for Oura’s CVA algorithm to record an individual baseline for each participant. The ergospirometry test will also be performed at the first visit. PWV measurement and randomization will be done on the second visit approximately two weeks after the first visit.

#### Cardiorespiratory fitness

To determine the intensity of the individually prescribed training program and the efficacy of the exercise intervention, ergospirometry tests will be carried out on a cycle ergometer (Ergoselect 4, Ergoline GmbH, Germany) at baseline and after 12 weeks for all three study groups. The test will start with a 1-minute sitting period in the saddle followed by a 3-minute warm-up with 0 watts (W). After that the workload will increase gradually according to an individualized ramp exercise protocol. The same individualized test protocol will be used at baseline and after 12 week intervention. The test will be supervised by an experienced exercise physiologist and participants will be verbally encouraged to continue until exhaustion. Respiratory gas exchange including ventilation and VO_2_max will be measured by the breath-by-breath method (Vyntus CPX, Vyaire Medical, USA). Electrocardiography will be recorded (Cardiosoft, USA) throughout the baseline exercise test.

#### Pulse wave velocity and pulse wave analysis

Carotid-femoral pulse wave velocity (cfPWV) measurements will be conducted in the morning following an overnight fast, during which participants are instructed to abstain from both caffeine and smoking. Measurements will be performed non-invasively using the Complior Analyze system (Alam Medical, France; Software v1.9.5), which is considered a non-invasive Gold Standard device for assessing arterial stiffness [11].

Following a 5-minute rest period in the supine position, the carotid pulse in the neck and the femoral pulse in the groin will be palpated. A small mark will be made at the site of the femoral pulse with a marker pen to ensure accurate distance measurement. The surface distance between the carotid and femoral sites will be measured using a flexible infant measuring device (Kern MSC 100, KERN & SOHN GmbH, Germany) and subsequently recorded in the Complior software. The device was selected because it measures straight-line distance independent of body shapes such as particularly large bellies and/or breasts in obese individuals. We only measured the distance between the carotid and femoral sites during the first visit to avoid introducing additional within-person measurement error [12].

A tonometric pulse sensor clamp will then be placed at the carotid artery site. The waveform morphology and signal quality will be visually inspected in the Complior software, and the measurement will proceed only when the signal is deemed acceptable. Subsequently, a finger-type sensor will be applied to the femoral pulse site. Pulse waveforms from both locations will be acquired simultaneously, and recordings will be accepted once the software indicates sufficient signal quality (≥95%). Each cfPWV measurement will be repeated at least three times to ensure reliability, and the results will be averaged for analysis.

Pulse wave analysis (PWA) will be performed non-invasively from the brachial artery using a cuff integrated in the same device. During the entire measurement protocol, participants will also wear an Oura Ring, enabling concurrent assessment of additional physiological parameters.

#### Other assessments

Standard demographics (age, sex, ethnic background according to United Nations Standard country or area codes for statistical use (M49)) were collected from all participants, and skin phototype was evaluated according to standard Fitzpatrick Scale. These are routinely collected for Oura’s internal PPG signal quality control to ensure that outputs remain consistent across various skin tones. In addition, body composition was determined by bioelectrical impedance (Inbody 720 body composition analyser, USA) in standing position after an overnight fast. Height was measured in a standing position. Female participants were also asked about their menstrual cycle phase, as it affects several of their composite scores and PWV measured by Oura Ring. Previous physical activity levels were assessed using the modified version of the short form of the International Physical Activity Questionnaire (IPAQ-SF). IPAQ is a set of questionnaires designed to assess physical activity levels in individuals aged 18 to 65 years [13]. All participants will also be asked to complete questionnaires at the beginning and end of the study to assess experiences of body composition measurement (RM4-FM), and exercise motivation and self-efficacy for exercising (Exercise Self-Efficacy Scale: ESES) in electronic form (Attachments labelled “RM4-FM” and “ESES”) made by using survey administration software (Google Forms).

#### Post Study Interviews

A random subgroup from each study group will be invited to a one-hour online qualitative interview at the end of the study. The aim of the interview study is to conduct a behavioral process evaluation of the exercise intervention to better understand how participants perceive changes in exercise habits and the use of Oura Ring to support exercise guidelines in each group. The interviews will be recorded and transcribed verbatim. About 8 participants from each group (N = 24) will be recruited for the interviews by invitation as they exit the main study. The study manager invited participants who had sufficient technological capability to take part in online interviews and ensured roughly equal numbers were invited from each study arm. Interactive tools will be used in the interviews to map experiences and general well-being paths during the study period. Participants will be asked to draw/point out their own experience and well-being path using these tools.

### Randomization

After the baseline measurements, the participants were randomized by permuted block randomization based on computer-generated random numbers. The block size was fixed to contain 9 participants: 3 to Control, 3 to HIIT and 3 to AI-based coaching groups. The participants were re-informed of the study design and procedures at the time of randomization. Study participants were told that we study methods for reducing arterial stiffness during a 12-week intervention. The randomization procedure involved participants choosing one of nine identically sealed opaque envelopes, which were in a mixed order containing the group assignment. The staff member who generated the random numbers and filled the sealed envelopes was independent of the individual who ran the randomization meeting, ensuring the allocation sequence was concealed until a participant chose an envelope.

The complete randomization list will be stored electronically on a secure, access-restricted private server. Access is limited to the independent researcher and the designated data manager, and all access events will be automatically logged. For allocation or other study-specific purposes, separate working files containing only the minimum necessary information are generated.

After the randomization, study participants were advised not to discuss issues related to group allocation during the assessments. All baseline data were collected prior to the randomization to prevent participants and study personnel from being biased due to group assignment. Outcome assessors will be blinded to the participants’ group assignment. The analysts conducting the primary analysis will be blinded to group codes until the intention-to-treat analysis is complete

### Interventions

Participants are instructed to maintain their usual diet, medication and lifestyle (other than the assigned exercise/control condition) throughout the 12-week intervention. Starting new structured exercise programs or using other lifestyle-tracking devices is prohibited.

The control group will be instructed to maintain their physical activity unchanged during the study. Leisure time physical activity at baseline as well as during the intervention period will be assessed by Oura Ring in all three study groups.

The HIIT instructor had the opportunity to motivate the participants in connection with the twice-weekly training sessions, which can be assumed to have improved adherence. The implementation of the AI coach was instructed in detail at the beginning of the intervention. Participants in all groups were able to contact the research staff at any time if they wished

#### High-intensity interval training (HIIT)

The exercise intervention started immediately after the second measurement visit. The HIIT group will follow a high-intensity interval training protocol on a cycle ergometer. In detail, the concept of HIIT involves repeated bouts of exercise at an intensity of 85% of W_max4_ interspersed by recovery periods based on a baseline ergospirometry test (Table 1). W_max4_ refers to the hypothetical workload sustainable for 4 min [14]. While being hypothetical, W_max4_ is useful for defining the presumably optimal intensity for work intervals at HIIT. The intervention group will perform HIIT sessions involving 5 bouts of 2–4 minutes work intervals (at 85% of W_max4_) interspersed by 3 minutes of active recovery (at 20% of W_max4_) period twice per week on non-consecutive days for 12 weeks. Each work interval will be 2 minutes long in the first week with 5 seconds added to each exercise session (i.e. 10 seconds per week) so that HIIT intervals will be 4 minutes long by week 12. If the participant fails to complete the work interval, they will still be instructed to continue the exercise session by starting the following scheduled work interval(s) after the recovery period(s). At the beginning of the intervention period, each HIIT session will last approximately 40 minutes, including warm-up (at 30% of W_max4_), recovery periods and cool-down (at 20% of W_max4_). At the end of the intervention period, each HIIT session will last approximately 50 minutes. HIIT will be conducted in a group of 1-4 participants closely supervised by the KuLTu personnel or students in health sciences adequately trained for the task.

**Table 1.** The exercise training program in the high-intensity interval training group. W_max4_ refers to the hypothetical workload sustainable for 4 minutes.

| Study months | 0–1 | 1–2 | 2–3 |
| --- | --- | --- | --- |
| *High-intensity interval training* |  |  |  |
| Duration of warm-up, min | 5 | 5 | 5 |
| High-intensity interval training frequency/week | 2 | 2 | 2 |
| Intensity of work phase, % $W_{\max4}$ | 85 | 85 | 85 |
| Intensity of recovery phase, % $W_{\max4}$ | 20 | 20 | 20 |
| Interval sets, number | 5 | 5 | 5 |
| Duration of the interval, seconds | 120-160 | 160-200 | 200-240 |
| Recovery between sets, minutes | 3 | 3 | 3 |
| Intensity of warm-up, % $W_{\max4}$ | 30 | 30 | 30 |
| Intensity of cool-down, % $W_{\max4}$ | 20 | 20 | 20 |
| Duration of cool-down, min | 5 | 5 | 5 |
| *Additionally home-based aerobic training* |  |  |  |
| Duration, min/week | 150-300 | 150-300 | 150-300 |

Additionally, an exercise training program will be prescribed for each participant in the HIIT group consisting of home-based low- to moderate-intensity aerobic exercises. An overall weekly goal, including supervised exercise, will be at least 150-300 minutes of aerobic exercise e.g. walking, swimming and cycling (Table 1) following international guidelines [10].

#### AI-based coach for steady-state aerobic training

KuLTu serves as the onsite study center responsible for participant recruitment, baseline and follow-up measurements, supervised HIIT sessions, and the initial setup of study devices and applications. All subsequent participant interaction with the AI system occurs remotely through the mobile application.

The AI-based coaching condition started immediately after the randomization when participants in this group were given access to an additional AI coaching feature, unavailable to participants in the other study groups. The clinical trial will use the production version of the AI coach with the addition of a prompt to focus on encouraging moderate, steady-state exercises 2-3 hours per week, also referred to as zone 2 cardio training.

The AI-based coaching feature and its supporting infrastructure are hosted offsite within Oura’s secure cloud environment. Physiological data captured by Oura Ring are transferred automatically to Oura’s cloud servers via the participant’s smartphone, processed to generate individualized insights, and returned to the participant through the app interface. Oura maintains the offsite computational resources, data-storage systems, and model-update processes.

The AI coach uses Large Language Models (LLM) to provide personalized health behavior coaching for users. The feature will primarily be a chat interface where the user can ask questions about their health data (e.g. Q: “How were my sleep scores last night?”; A: “Last night, it seems you had about 7 hours of sleep with some restlessness. Your Sleep Score was 83”) and discuss non-clinical behavior change (e.g. Q: “I’ve been thinking I’d like to do more stretching”; A: “Flexibility and mobility are key. Consider focusing on stretches that target the hips, groin, hamstrings, and lower back.”). This creates a feedback-loop of observing health data, making behavioral changes and tracking potential changes. AI coach will have access to the user’s health data and can create a memory out of things the user tells them about their life and preferences, which it can use to make non-clinical recommendations to the user that are adapted to their unique personal data, circumstances and preferences. The user has full access to, and ability to delete, the memories created by the feature. The foundation model vendor does not have direct access to user data, only through an application programming interface, which means user data cannot be used by the vendor for LLM training.

##### AI algorithm version and update management

The AI-based coaching feature used in this trial will be *Oura Advisor*, integrated within the Oura App. Participants will access the frozen production build active between October 2024 and May 2025. During the study period, Oura is expected to release routine updates to the consumer app environment. Engineering records will later be reviewed to determine which of these deployments affect the Advisor component accessible to participants.

All updates are expected to primarily involve interface refinements, bug fixes, or adjustments to data-fetch and display systems. No modifications will be made to the underlying large-language-model (LLM) weights or to the recommendation algorithms. The prompt configuration governing the Advisor’s behavior will remain fixed throughout the study period. Planned updates may include minor improvements to how data are structured or presented to the model, but such changes will not alter the substantive content or logic of recommendations.

A version-control log will be maintained by Oura’s engineering team. This log will include build identifiers, deployment dates, and a short description of each update’s functional scope.

##### Input-data-level inclusion and exclusion criteria

The AI-based coaching system processes physiological and behavioral input data generated by Oura Ring and Oura App to generate personalized feedback for participants. Data are accepted by the AI system only when sensor and synchronization quality meet predefined thresholds:

- PPG and motion signals meet Oura’s internal quality-control thresholds (signal-quality ≥ 95 % for nightly HRV; adequate wear time ≥ 20 h/day on ≥ 5 days/week);
- The ring and app remain synchronized via Bluetooth and cloud connection; and
- User-initiated text interactions occur within the study period and language supported by the system.

Data are excluded when signal quality is insufficient, the ring is not worn for ≥ 3 consecutive days, the app is uninstalled, or the participant withdraws consent for data use. These technical criteria ensure that the AI receives input consistent with the data distributions on which its algorithms were developed and validated.

##### Acquisition and selection of input data

Physiological inputs are acquired passively by Oura Ring, which records photoplethysmography (PPG), accelerometer, and temperature data. These data are encrypted, transferred via Bluetooth to the participant’s smartphone, and uploaded automatically to Oura’s secure cloud servers. The AI coach accesses processed daily summaries (e.g., sleep duration, HRV, resting heart rate, activity rate) through the same internal data pipeline used for the standard Oura App features. Only validated, successfully uploaded data are accessible to the AI coach; no raw sensor streams are analyzed locally on the phone. In addition, participants may provide voluntary contextual information (e.g., preferences, barriers, exercise intentions) through text chat. These text exchanges are stored in the user’s encrypted account and accessed by the AI for personalization.

##### Handling of poor-quality or unavailable input data

Data-quality assessment is performed automatically within Oura’s cloud infrastructure prior to any AI processing. PPG and motion signals are continuously quality-checked; segments that fail to meet internal thresholds (e.g., signal-quality < 95 % or missing synchronization for > 24 h) are flagged as invalid. When insufficient data are available, the AI coach defaults to generalized guidance based on the user’s recent behavioral trends rather than producing data-specific recommendations. Missing or poor-quality periods are surfaced to participants through standard Oura notifications encouraging ring wear or device synchronization. All raw and flagged data remain in the research database for completeness but are excluded from AI-generated personalization.

##### Human–AI interaction in input handling

No manual preprocessing or expert adjudication of physiological data occurs before AI processing; all data selection and quality control are automated within Oura’s pipeline. Human involvement is limited to participant setup and troubleshooting at baseline visits. Participants themselves interact with the AI coach through natural-language chat in plain English. No technical or clinical expertise is required beyond basic smartphone literacy. Research personnel at KuLTu oversee adherence and safety but do not handle raw sensor data or intervene in the AI’s preprocessing.

##### Output of the AI intervention

The AI coach produces text-based conversational outputs that deliver personalized, non-clinical behavior-change guidance. Outputs include daily or weekly motivational messages, reflections on progress, and specific suggestions for moderate steady-state aerobic exercise (“zone 2” training) based on recent Oura metrics. Responses appear within the in-app chat interface and are visible only to the participant. No diagnostic or prescriptive medical advice is generated, and no automated alerts are sent to researchers or clinicians.

##### Role of AI output in decision-making

The AI output contributes to participant decision-making by guiding self-directed exercise behavior during the 12-week intervention. Participants may choose to follow or disregard the AI’s suggestions; no clinical decisions or safety monitoring depend on the AI. Research staff and clinicians do not view or act on AI messages. Thus, the AI serves as a behavioral-support tool within a preventive-health context rather than a component of clinical care.

##### Performance Error Identification

The AI-based coaching feature will operate as a stable, production-level system throughout the intervention period with the addition of the study specific prompt section, instructing the focus on steady-state cardio. The research team will not perform real-time monitoring or iterative modification of the algorithm or its outputs during the trial. Routine system oversight, including automated logging of software bugs, connectivity issues, and latency events, will be conducted by Oura’s engineering team as part of the company’s standard production maintenance, rather than as a trial-specific activity.

Following completion of the 12-week intervention, Oura and the study team will conduct a retrospective assessment of Advisor chat performance to identify any technical issues or irregularities that may have affected participant interactions. The retrospective analysis will focus on software reliability, message-delivery consistency, and adherence to the intended behavioral-support scope. A summary of this evaluation will be included in the final study report.

##### Access to the AI Intervention and Code

The underlying model architecture, prompting framework, and source code will remain confidential and will not be publicly available due to commercial and data-security restrictions. The version deployed for this study will be identical to the production version available to Oura Members, with the exception of configuration settings that specify the study’s focus on steady-state aerobic training.

For research transparency, upon reasonable request Oura may consider providing the investigators with more detailed descriptions summarizing the Advisor’s operational scope. Technical details of any algorithms, data inputs and outputs, quality-control mechanisms, and version history for the duration of the study are proprietary material. External access to the AI codebase or model weights will not be granted.

### Statistical Analysis

The statistical analysis for this study will employ a comprehensive approach that combines both frequentist and Bayesian frameworks. This dual-methodology is chosen due to the inherent challenges of this trial, including the known variability of arterial stiffness, and the complexities of adherence to a behavioral intervention. We postulate that relying on a single analytical approach would provide an incomplete picture of the evidence.

For our frequentist analysis, instead of relying on a strict dichotomization of results based on a single p-value threshold, we will interpret p-values as a continuous measure of evidence against the null hypothesis. The p-values will be reported in full, and the strength of the evidence will be discussed in context with effect sizes, confidence intervals, and the clinical significance of the findings. This approach aligns with modern statistical recommendations that discourage the oversimplification of results into "statistically significant" or "not significant."

In parallel, we will conduct a Bayesian analysis to provide a more nuanced understanding of our data. This approach allows us to quantify the probability of an effect existing and to incorporate prior knowledge about the intervention’s likely impact. We will specify our priors transparently and, where appropriate, conduct sensitivity analyses to assess the robustness of our conclusions to different prior assumptions. The results from the Bayesian analysis, including posterior probabilities and credible intervals, will complement the frequentist results by offering a more direct assessment of the evidence for or against our hypotheses.

We commit to reporting the results from both frequentist and Bayesian analyses openly and without bias, regardless of whether they lead to similar or different conclusions. This preregistered plan is intended to enhance transparency, mitigate the risk of selective reporting, and provide the most complete and robust interpretation of our study’s findings. We recognize that lifestyle intervention trials are challenging, especially when the outcome variable is highly heterogenous. Given our study’s limited size and somewhat complex design and duration, we can realistically expect to detect clinically small differences between the groups, and therefore all conclusions will be interpreted as exploratory, warranting larger and more specific trials for more robust conclusions.

### Power calculations

Sample size for this study is based on detecting a change in PWV as measured by the Gold Standard reference device (Complior device, Alam Medical, France). We also leverage data from the Pilot Trial. Previous literature has shown that 4-8 weeks of aerobic exercise can reduce PWV by −0.35 (−0.68, −0.02) m/s; and a 9-16 week intervention by −0.69 (−1.13, −0.25) m/s [6]. This outcome is heavily dependent on changes in VO_2_max; for example, a less than 10% improvement will only result in −0.4 (−0.52, −0.28) m/s change in PWV, whereas >20% improvement will result in −1.72 (−2.39, −1.04) m/s change. Based on the pilot trial, we estimate that the conservative effect (−0.4 m/s) is more realistic, and this is selected as the expected effect size. We also know from the Pilot that the standard deviation in PWV is quite large relative to the expected effect size. By sampling the Pilot data, we obtain a SD of 1.4 m/s for the 30-65-year age group. Using these numbers in standard power calculations [15] result in N = 193 per group, assuming 80% power at alpha-level 0.05 (two-sided test). Based on the Pilot, we also know that the correlation in pre- and post-intervention PWV measurements in the control group is high (R = 0.86) and it’s mostly explained by age, sex, BMI and blood pressure. While the results will be analyzed using a mixed linear model (see below), for which there is no formal power calculation protocol, we leverage the sample size calculations developed by Borm et al. [16] for analysis of covariance (ANCOVA) and adjust the sample size with (1-⍴^2^), where ⍴ is the correlation coefficient for pre- and post-intervention outcome variables. This yields a sample size of 50 per group, and to account for attrition, a 10% increase is considered sufficient, leading to a final number of 55 per group.

#### Frequentist analysis plan

The primary frequentist analysis will be based on the Intention-to-Treat (ITT) framework, defined as all randomized participants included in the groups to which they were initially assigned, regardless of adherence or withdrawal. The mixed linear model (see below), which inherently accommodates longitudinal data with missing time points under the assumption of Missing At Random (MAR), will be the primary method used to analyze this ITT population. The ITT analysis provides an unbiased estimate of the intervention’s effectiveness under real-world conditions. However, we expect highly motivated participants to engage the most in their assigned interventions and thus receive the most benefit. This will be investigated in mediator analyses. It is also possible that the intervention works differently for certain people (e.g. men vs. women) and these will be studied as moderators. Both mediator and moderator analyses are described below.

The primary analysis will be based on a mixed linear model, which is an appropriate method for analyzing repeated measures data while accounting for individual-level variability. This model will be used to assess the effectiveness of the interventions and compare the mean post-intervention cfPWV and VO_2_max across the three study groups.

The difference between the AI group and Oura Ring only -group will be considered the primary comparison, while differences across other group pairs will be exploratory. We will assume that any missing data is MAR. The results will be presented with full, numeric p-values, mean differences, and 95% confidence intervals, avoiding an arbitrary (e.g. p = 0.05) dichotomy into statistically “significant” and “insignificant.”

To evaluate how strongly candidate mediators account for the intervention–outcome association, each mediator will be entered sequentially into the previously specified linear mixed model, and the resulting change in the intervention regression coefficients will be examined. The proportion of attenuation attributable to each mediator will be calculated as: (1 − [regression coefficient adjusted for mediator / regression coefficient adjusted for determinant]) × 100%. In parallel, we will assess the incremental predictive value contributed by the intervention effects beyond their presumed mediators. The mediation analyses will be conducted in the full participant population across all randomization groups.

In addition to mediator analyses, we will evaluate pre-specified moderators (e.g., sex, age, baseline cfPWV, cardiorespiratory fitness) by extending the primary linear mixed model to include the group × time × moderator interaction. For binary moderators (e.g., sex), inference will focus on the group × time × sex term; for continuous moderators, variables will be mean-centered and effects probed at interpretable values (e.g., mean ±1 SD) via model-based marginal means and simple-slope contrasts. When an interaction shows evidence of effect modification, we will present stratified (subgroup) estimates and contrasts within moderator levels for interpretation.

These secondary analyses are crucial for exploring potential effects that may be obscured by the ITT analysis, particularly if adherence is low. They will be clearly identified as exploratory and will complement the primary ITT analysis.

#### Bayesian analysis plan

In parallel with the frequentist analysis, we will conduct a Bayesian analysis for the primary and key secondary outcomes (cfPWV and VO_2_max change). The Bayesian model will mirror the mixed linear model used in the frequentist framework to ensure direct comparability of results; specifically, we will include a random intercept at the individual level, fixed effects for the three treatment groups, and covariate adjustment for baseline outcome, age, sex, BMI, and systolic blood pressure. By aligning the model structure, any differences between approaches will reflect the inferential framework (use of priors and posterior probabilities) rather than model specification.

Priors will be specified transparently and chosen to balance prior knowledge with appropriate uncertainty. For the primary treatment effect on cfPWV, we will use a Normal prior with mean −0.4 m/s and standard deviation 0.5 m/s, centered on our expectation derived from an unpublished pilot study, while allowing the posterior to span both benefit and no effect. For VO_2_max, we will use a weakly informative Normal prior with mean 0 and standard deviation 5 mL·kg^-1^·min^-1^, covering plausible ±10 mL·kg^-1^·min^-1^ changes. Covariate effects will receive Normal(0, 10) priors to mirror the unpenalized adjustments in the frequentist model. Variance parameters, including the standard deviations of the individual random intercepts and the residual error, will be given half-Normal(0, 5) priors to provide mild regularization of variability estimates.

The likelihood assumes normally distributed post-intervention outcomes with participant-specific random intercepts and a common residual variance. We will sample from the joint posterior distribution of all parameters and assess convergence using split-chain diagnostics (e.g., agreement across chains) alongside effective sample size thresholds to ensure stable estimation. Posterior summaries will be presented as means and 95% credible intervals for each treatment contrast (AI vs. control; HIIT vs. control; AI vs. HIIT), providing directly interpretable intervals for the magnitude and direction of effects.

To aid clinical interpretation, we will also report posterior probabilities for prespecified thresholds of meaningful benefit; for example, the probability that the AI versus control effect on cfPWV is less than −0.2 m/s. Finally, we will evaluate robustness to prior assumptions by repeating the primary cfPWV analysis under a skeptical prior (Normal mean 0, SD 0.3 m/s) and an enthusiastic prior (Normal mean −0.6, SD 0.3 m/s). These sensitivity analyses will clarify how much the conclusions depend on the choice of prior while preserving the underlying model structure.

#### Process Evaluation

The study will include a process evaluation to examine (i) the implementation of the intervention study, (ii) context that may have influenced the study and its findings, and (iii) the mechanisms of the intervention’s effects.

Process evaluations are well-established in public health and health service research, where they are considered essential to understanding how complex interventions achieve their effects [17,18]. “Complex interventions” are health interventions that have many potential “active ingredients.” A complex intervention combines different components in a whole that is more than the sum of its parts. While process evaluation is commonplace in public health and service settings, its purpose and aims are equally relevant for evaluating any intervention in which multiple factors and influences may shape trial outcomes. In this study, the interventions are complex in that they integrate structured exercise, technology-supported self-monitoring, AI delivered behavioral support, and participants’ own contextual circumstances.

##### (i) Implementation

Implementation will be assessed to understand how delivery of the interventions was achieved, what was delivered, and to whom. Trial records will be used to monitor numbers randomized, adherence to baseline and follow-up assessments, and attrition across groups. Intervention delivery will be captured through adherence to HIIT sessions and the number and frequency of Advisor interactions in the AI coach group.

##### (ii) Context

Context will be explored to identify external and personal factors that influenced participant engagement with the interventions. Post-study interviews with a subsample of approximately 8 participants from each study arm will investigate participants’ life circumstances (e.g., health, family or work demands, seasonality) that may have shaped their ability to participate. In addition, the technological environment will be considered: as the study is conducted with a commercial wearable and companion app, feature updates or design changes released during the trial may alter how participants interact with the intervention. Such changes will be documented and examined as contextual influences, while participants’ perceptions of these changes will be captured through the interviews.

##### iii) Mechanisms of impact

Mechanisms of impact will be examined to understand how participants engaged with and experienced the interventions, and how these contributed to perceived or actual changes in behavior. Post-study interviews will explore participants’ perceptions of changes in exercise habits, motivation, and use of Oura Ring and AI coach.

Questionnaire data from the RM4-FM (body composition measurement experiences) and the Exercise Self-Efficacy Scale (ESES) will provide descriptive markers of participants’ experiences and self-efficacy across groups. In addition, Advisor chat logs will be summarized descriptively to identify common conversation topics, user trajectories, and pain points.

##### Qualitative process evaluation

Interview transcripts will be analyzed using reflexive thematic analysis, supported by artificial intelligence (AI) tools such as LLM for coding assistance, clustering, and retrieval of exemplar quotes [19-21]. Final theme development and interpretation will remain human-led. Coding will proceed iteratively, with codes and themes discussed within the research team to ensure reflexivity and consistency. A subset of transcripts will be double-coded to enhance credibility. Interactive visual tools used during interviews will also generate participant-drawn trajectories of experience and well-being, which will be incorporated into the analysis.

##### Quantitative descriptive process evaluation

Questionnaire data will be summarized descriptively to provide additional context for interpreting study outcomes. RM4-FM and ESES scores will be reported as means and standard deviations (or medians and interquartile ranges if non-normally distributed) at baseline and 12 weeks for each group. Within-group descriptive changes over time will be summarized. Engagement metrics, including Oura App analytics, and frequency and length of Advisor chat interactions, will be reported descriptively. Advisor chat analytics will be generated using Nebuly, a user analytics platform for LLM applications that automatically captures conversational interactions while anonymizing personal information; these data will provide aggregate summaries such as the number and length of conversations, common user intents, and interaction drop-off points. These data are not intended for mediation or moderation analyses, but to contextualize participants’ reported experiences of the study and interventions.

### Ethical and safety considerations

Oura Ring is strictly for consumer use only and not intended to be a medical device. The study plan has been approved by The Regional Medical Research Ethics Committee of Eastern Finland Collaborative Area. The study aims, design and progression and the content of study visits will be discussed with the study participants in detail. They will also receive written information on these aspects. The participants give their written consent to participate in the study. They have every right to discontinue the study at any point without providing a reason.

There will be no significant risks to participate in the present study. An experienced exercise physiologist will supervise each ergospirometry test, and each HIIT session will be supervised by students in health sciences or somebody from the KuLTu personnel adequately trained for the task. Personnel have up-to-date first aid skills and a defibrillator will be available during the ergospirometry tests and also during supervised HIIT sessions. The well-being of the subjects will be followed during the study visits.

Participants are invited to participate in the process evaluation interviews voluntarily, but they can withdraw their consent at any stage. The qualitative interview methods used in this study are typical of qualitative research and are widely used. The researchers conducting the study have several years of experience in conducting similar studies and are thoroughly familiar with the methods used.

Interviews that address health and personal life topics always carry the possibility that questions may touch on sensitive and potentially emotional topics for the interviewees. Interviewees are not obliged to answer any questions they do not wish to answer and will be reminded of this verbally during the interview.

Due to the low-risk nature of the exercise intervention and the non-medical classification of the device features, no formal, independent Data Monitoring Committee (DMC) is considered necessary. Any participant who suffers harm from trial participation will be provided with necessary ancillary care and follow-up by the Kuopio University Hospital. There is no provision for financial compensation other than that mandated by local research ethics guidelines.

Any significant protocol amendments (e.g., changes to eligibility, outcomes, or analysis) will be communicated to The Regional Medical Research Ethics Committee of Eastern Finland Collaborative Area and the ClinicalTrials.gov registry and will be detailed in the final study report.

### The anonymity and storage of the data

All data will be coded so that no one except the study nurse and the responsible researcher at KuLTu will be able to link the samples to the individual participant. The data is coded with a study ID that is used in all measurements of the participant. All of the collected data is confidential and will not be given to anyone without permission. Data are stored for the duration of the study (15 years, as included in the approved IRB application) and then destroyed.

All mentions of names or places will be removed from anonymized interview transcripts. Interview data will be coded with pseudonyms and grouped according to the intervention groups. The interview recordings will be destroyed after transcription is complete. Transcripts will be retained for the duration of the study (15 years) and then destroyed.

AI coach has undergone legal review with the Oura legal team and sufficient protections have been implemented, including vendor security questionnaires and contractual agreements, to ensure any service providers process personal data to the same stringent standards as Oura.

The final, de-identified participant-level dataset and the statistical code will be made publicly available via a recognized repository upon publication of the main results manuscript, in accordance with the journal’s policies, pending explicit, additional permission from the participants.

The final trial dataset (coded, de-identified) will be accessible only to the authors responsible for data analysis (HP, SS, DC and PO). There are no contractual agreements between institutions that limit access to the anonymized final trial dataset for these investigators

## Results and publication

The recruitment for the study began in October 2024 and will conclude once 165 men and women have been recruited. Data collection for the study is scheduled to conclude in March 2026. At the time of writing this protocol, three participants have withdrawn from the study during the intervention phase. Data management is ongoing.

The peer-reviewed study protocol will be accessible via the journal’s website and the study’s site at clinicaltrials.gov. A separate Statistical Analysis Plan (SAP) is not currently planned as all necessary details are included within this protocol.

## Discussion

Rationale for the study design is to determine what are the effects of adopting a wearable device with built-in AI guidance and exercise on vascular aging/stiffness and cardiorespiratory fitness. Based on existing literature, improving aerobic fitness leads to a reduction in PWV, but little is known about the possibly differing effects of different exercise modalities. For example, is HIIT equivalent to steady-state aerobic training? It is also widely recognized that adherence to regular exercise can be a major challenge for many people. Oura can provide continuous user engagement through a phone app, which displays the user’s personal data. The same app can be used to provide AI-based coaching tailored to the user, which may also improve CVA.

With any wearable technology, a critical objective is finding the “active ingredient” - is it simply the purchase of a new device and thus becoming more mindful of one’s behavior, or is it some specific feature of the device/app? To disentangle some of the correlated effects, we will use a control group that receives the ring itself with all the available data except CVA. Using this as a comparison, we can see if the added exercise makes a difference compared to just owning an Oura Ring.

Can an AI coach cause Oura users to improve their VO_2_max equally to supervised HIIT exercise? This is a critical question, because it can be safely assumed that long-term adherence to a strenuous HIIT protocol will be unsustainable for most people. This is why a flexible, continuously accessible AI coach could prove to be an invaluable tool for maintaining regular exercise routines.

Participant adherence to the study thus far has been excellent, with only three participants withdrawing during the intervention phase.

Gold standard methodologies are employed to assess VO_2_max and PWV. We also objectively measure data on participants’ leisure-time physical activity using Oura Ring. The HIIT protocol is individually tailored and all exercise sessions are rigorously monitored by professionals to ensure protocol adherence and participant safety.

In addition to these methodological strengths, the embedded process evaluation will provide critical insights into how the interventions were delivered and experienced. This is especially important for trials of complex interventions such as supervised HIIT and AI-based coaching, where multiple interacting components and contextual influences may shape outcomes. By examining implementation, participant context, and mechanisms of impact through a combination of interviews, questionnaires, engagement metrics, and Advisor chat analytics, we will be able to better interpret whether observed effects reflect the intervention design, the fidelity of its delivery, or external factors such as app updates or participants’ personal life circumstances. Process evaluation findings will also help to identify the “active ingredients” of engagement with Oura Ring and the AI coach, complementing the trial’s quantitative outcomes with explanatory insights that can inform both scientific interpretation and future product development.

### Limitations

By default, the AI coach operates in English, which is a non-native language to the study participants. However, they are able to ask questions in Finnish, too, in which case the responses are also provided in Finnish. These outputs are instructed to be interpreted as clarifying the English outputs, since the Finnish outputs are based on much more limited LLM training data. None of the participants randomly assigned to the AI group has reported the use of English in the application as a barrier that would have discouraged them from participating in the study.

## Conclusion

In this RCT, we will study the effects of supervised HIIT and AI-based coaching on reducing arterial stiffness measured by a Gold Standard reference device and by Oura Ring. The results are expected to be valuable, as they contribute to the understanding of how commercially available smart ring technology and adopting AI-guided physical activity routines may support vascular health.

## Supporting information

Appendix 2 (SPIRIT-AI Checklist)

Appendix 1 (WHO structured abstract))

## Data Availability

All data produced in the present study are available upon reasonable request to the authors; pending additional consent from the participants

## Acknowledgments

This study was funded by Oura Health Oy (contact information for the sponsor; corresponding author PO). We would like to thank the staff involved in the study and all the participants. Gemini 2.5 was used for writing assistance of this manuscript, but not for study design, data collection or analysis.

## Conflicts of interest

SS, JK, AR, HS, DC and PO declare competing financial interests in the form of ongoing salaried employment and employee equity at Oura Health Oy (Oulu, Finland) and MdZ at Ouraring Inc (San Francisco, CA, United States) or any of their affiliates. While the authors have a professional affiliation with the company, they have conducted this research with the highest standards of scientific integrity and transparency. HP, SH, PK and KS declare no competing interests.

## Authors’ contributions

HP, SS, JK, AR, SH, HS, DC, MdZ, KS, PK and PO designed the study protocol and contributed to the coordination and implementation of the research. HP, SS, DC, MdZ and PO wrote the manuscript. KS assisted in developing research measures and collecting data. All authors participated in drafting the manuscript and have read and approved the final version. No professional writers outside the authors listed in this manuscript will be used.

## Abbreviations

AI: artificial intelligence
CVA: cardiovascular age
HIIT: high-intensity interval training
PWV: pulse wave velocity
RCT: Randomized controlled trial

## Notes

### Competing Interest Statement

All authors have completed the ICMJE uniform disclosure form at www.icmje.org/coi_disclosure.pdf and declare: SS, JK, AR, HS, DC and PO declare competing financial interests in the form of ongoing salaried employment and employee equity at Oura Health Oy (Oulu, Finland) and MdZ at Ouraring Inc (San Francisco, CA, United States) or any of their affiliates. HP, SH, PK and KS declare no financial relationships with any organizations that might have an interest in the submitted work in the previous three years. All authors declare no other relationships or activities that could appear to have influenced the submitted work.

### Clinical Trial

NCT06644014

### Funding Statement

This study was funded by Oura Health Oy (Oulu, Finland)

### Author Declarations

Approval granted by Regional Medical Research Ethics Committee of Eastern Finland Collaborative Area

