## Appendix 2 (SPIRIT-AI Checklist) for "The Effects of AI-Guided Exercise and a Smart Ring on Arterial Stiffness (GONDOR-AS): protocol for a randomized controlled trial"

### Appendix 2. SPIRIT-AI Checklist: Recommended items to address in a protocol and related documents for clinical trials evaluating AI interventions

| Section | | SPIRIT 2013 Item[a] | SPIRIT-AI Item | | Addressed on Page No [b] |
|---|---|---|---|---|---|
| **Administrative Information** | | | | | |
| Title | 1 | Descriptive title identifying the study design, population, interventions, and, if applicable, trial acronym | SPIRIT-AI 1(i) Elaboration | Indicate that the intervention involves artificial intelligence / machine learning and specify the type of model. | P 1-3, 10-13 |
| | | | SPIRIT-AI 1(ii) Elaboration | Specify the intended use of the AI intervention. | P 1-4 |
| Trial registration | 2a | Trial identifier and registry name. If not yet registered, name of intended registry | | | P 1, 4 |
| | 2b | All items from the World Health Organization Trial Registration Data Set | | | Appendix 2 |
| Protocol version | 3 | Date and version identifier | | | P 1 |
| Funding | 4 | Sources and types of financial, material, and other support | | | P23 |
| Roles and responsibilities | 5a | Names, affiliations, and roles of protocol contributors | | | P 1 |
| | 5b | Name and contact information for the trial sponsor | | | P 23 |
| | 5c | Role of study sponsor and funders, if any, in study design; collection, management, analysis, and interpretation of data; writing of the report; and the decision to submit the report for publication, including whether they will have ultimate authority over any of these activities | | | P 23 |
| | 5d | Composition, roles, and responsibilities of the coordinating centre, steering committee, endpoint adjudication committee, data management team, and other individuals or groups overseeing the trial, if applicable (see Item 21a for data monitoring committee) | | | P 20, 23 |
| **Introduction** | | | | | |
| Background and rationale | 6a | Description of research question and justification for undertaking the trial, including summary of relevant studies (published and unpublished) examining benefits and harms for each intervention | SPIRIT-AI 6a (i) Extension | Explain the intended use of the AI intervention in the context of the clinical pathway, including its purpose and its intended users (e.g. healthcare professionals, patients, public). | P 3-4 |
| | | | SPIRIT-AI 6a (ii) Extension | Describe any pre-existing evidence for the AI intervention. | P 3 |
| | 6b | Explanation for choice of comparators | | | P 4 |
| Objectives | 7 | Specific objectives or hypotheses | | | P 4 |

Cite as:

| | | | | | |
|---|---|---|---|---|---|
| Trial design | 8 | Description of trial design including type of trial (eg, parallel group, crossover, factorial, single group), allocation ratio, and framework (eg, superiority, equivalence, noninferiority, exploratory) | | | P 4 |
| Methods: Participants, Interventions and Outcomes | | | | | |
| Study setting | 9 | Description of study settings (eg, community clinic, academic hospital) and list of countries where data will be collected. Reference to where list of study sites can be obtained | SPIRIT-AI 9 Extension | Describe the onsite and offsite requirements needed to integrate the AI intervention into the trial setting. | P 10 |
| Eligibility criteria | 10 | Inclusion and exclusion criteria for participants. If applicable, eligibility criteria for study centres and individuals who will perform the interventions (eg, surgeons, psychotherapists) | SPIRIT-AI 10 (i) Elaboration | State the inclusion and exclusion criteria at the level of participants. | P 5 |
| | | | SPIRIT-AI 10 (ii) Extension | State the inclusion and exclusion criteria at the level of the input data. | P 11-12 |
| Interventions | 11a | Interventions for each group with sufficient detail to allow replication, including how and when they will be administered | SPIRIT-AI 11a (i) Extension | State which version of the AI algorithm will be used. | P 11 |
| | | | SPIRIT-AI 11a (ii) Extension | Specify the procedure for acquiring and selecting the input data for the AI intervention. | P 11-12 |
| | | | SPIRIT-AI 11a (iii) Extension | Specify the procedure for assessing and handling poor quality or unavailable input data. | P 12 |
| | | | SPIRIT-AI 11a (iv) Extension | Specify whether there is human-AI interaction in the handling of the input data, and what level of expertise is required for users. | P 12 |
| | | | SPIRIT-AI 11a (v) Extension | Specify the output of the AI intervention. | P 12-13 |
| | | | SPIRIT-AI 11a (vi) Extension | Explain the procedure for how the AI intervention's output will contribute to decision-making or other elements of clinical practice. | P 13 |
| | 11b | Criteria for discontinuing or modifying allocated interventions for a given trial participant (eg, drug dose change in response to harms, participant request, or improving/worsening disease) | | | P 5 |
| | 11c | Strategies to improve adherence to intervention protocols, and any procedures for monitoring adherence (eg, drug tablet return, laboratory tests) | | | P 8 |
| | 11d | Relevant concomitant care and interventions that are permitted or prohibited during the trial | | | P 8 |
| Outcomes | 12 | Primary, secondary, and other outcomes, including the specific measurement variable (eg, systolic blood pressure), analysis metric (eg, change from baseline, final value, time to event), method of aggregation (eg, median, proportion), and time point for each outcome. Explanation of the clinical relevance of chosen efficacy and harm outcomes is strongly recommended | | | P 2, 4 |
| Participant timeline | 13 | Time schedule of enrolment, interventions (including any run-ins and washouts), assessments, and visits for participants. A schematic diagram is highly recommended (see Figure) | | | P 5 |

| | | | | | |
|---|---|---|---|---|---|
| Sample size | 14 | Estimated number of participants needed to achieve study objectives and how it was determined, including clinical and statistical assumptions supporting any sample size calculations | | | P 15 |
| Recruitment | 15 | Strategies for achieving adequate participant enrolment to reach target sample size | | | P 4 |
| Methods: Assignment of Interventions (For Controlled Trials) | | | | | |
| Sequence generation | 16A | Method of generating the allocation sequence (eg, computer-generated random numbers), and list of any factors for stratification. To reduce predictability of a random sequence, details of any planned restriction (eg, blocking) should be provided in a separate document that is unavailable to those who enrol participants or assign interventions | | | P 7-8 |
| Allocation concealment mechanism | 16b | Mechanism of implementing the allocation sequence (eg, central telephone; sequentially numbered, opaque, sealed envelopes), describing any steps to conceal the sequence until interventions are assigned | | | P 8 |
| Implementation | 16c | Who will generate the allocation sequence, who will enrol participants, and who will assign participants to interventions | | | P 7-8 |
| Blinding (masking) | 17a | Who will be blinded after assignment to interventions (eg, trial participants, care providers, outcome assessors, data analysts), and how | | | P 8 |
| | 17b | If blinded, circumstances under which unblinding is permissible, and procedure for revealing a participant's allocated intervention during the trial | | | N/A |
| Methods: Data Collection, Management, And Analysis | | | | | |
| Data collection methods | 18a | Plans for assessment and collection of outcome, baseline, and other trial data, including any related processes to promote data quality (eg, duplicate measurements, training of assessors) and a description of study instruments (eg, questionnaires, laboratory tests) along with their reliability and validity, if known. Reference to where data collection forms can be found, if not in the protocol | | | P 5-7 |
| | 18b | Plans to promote participant retention and complete follow-up, including list of any outcome data to be collected for participants who discontinue or deviate from intervention protocols | | | P 5 |
| Data management | 19 | Plans for data entry, coding, security, and storage, including any related processes to promote data quality (eg, double data entry; range checks for data values). Reference to where details of data management procedures can be found, if not in the protocol | | | P 20-21 |
| Statistical methods | 20a | Statistical methods for analysing primary and secondary outcomes. Reference to where other details of the statistical analysis plan can be found, if not in the protocol | | | P 15-17 |
| | 20b | Methods for any additional analyses (eg, subgroup and adjusted analyses) | | | P 15-17 |

Cite as:

| | | | | | |
|---|---|---|---|---|---|
| | 20c | Definition of analysis population relating to protocol non-adherence (eg, as randomised analysis), and any statistical methods to handle missing data (eg, multiple imputation) | | | P 15 |
| Methods: Monitoring | | | | | |
| Data monitoring | 21a | Composition of data monitoring committee (DMC); summary of its role and reporting structure; statement of whether it is independent from the sponsor and competing interests; and reference to where further details about its charter can be found, if not in the protocol. Alternatively, an explanation of why a DMC is not needed | | | P 20 |
| | 21b | Description of any interim analyses and stopping guidelines, including who will have access to these interim results and make the final decision to terminate the trial | | | N/A |
| Harms | 22 | Plans for collecting, assessing, reporting, and managing solicited and spontaneously reported adverse events and other unintended effects of trial interventions or trial conduct | SPIRIT-AI 22 Extension | Specify any plans to identify and analyse performance errors. If there are no plans for this, explain why not. | P 13 |
| Auditing | 23 | Frequency and procedures for auditing trial conduct, if any, and whether the process will be independent from investigators and the sponsor | | | N/A |
| Ethics and Dissemination | | | | | |
| Research ethics approval | 24 | Plans for seeking research ethics committee/institutional review board (REC/IRB) approval | | | P 19 |
| Protocol amendments | 25 | Plans for communicating important protocol modifications (eg, changes to eligibility criteria, outcomes, analyses) to relevant parties (eg, investigators, REC/IRBs, trial participants, trial registries, journals, regulators) | | | P 20 |
| Consent or ascent | 26a | Who will obtain informed consent or assent from potential trial participants or authorised surrogates, and how (see Item 32) | | | P 19 |
| | 26b | Additional consent provisions for collection and use of participant data and biological specimens in ancillary studies, if applicable | | | P 19 |
| Confidentiality | 27 | How personal information about potential and enrolled participants will be collected, shared, and maintained in order to protect confidentiality before, during, and after the trial | | | P 20 |
| Declaration of interests | 28 | Financial and other competing interests for principal investigators for the overall trial and each study site | | | P 23 |
| Access to data | 29 | Statement of who will have access to the final trial dataset, and disclosure of contractual agreements that limit such access for investigators | SPIRIT-AI 29 Extension | State whether and how the AI intervention and/or its code can be accessed, including any restrictions to access or re-use. | P 13 |
| Ancillary and post-trial care | 30 | Provisions, if any, for ancillary and post-trial care, and for compensation to those who suffer harm from trial participation | | | P 20 |
| Dissemination policy | 31a | Plans for investigators and sponsor to communicate trial results to participants, healthcare professionals, the public, and other relevant groups (eg, via publication, reporting in results databases, or other data sharing arrangements), including any publication restrictions | | | P 21 |

Cite as:

| | | | | | |
|---|---|---|---|---|---|
| | 31b | Authorship eligibility guidelines and any intended use of professional writers | | | P 23 |
| | 31c | Plans, if any, for granting public access to the full protocol, participant-level dataset, and statistical code | | | P 20-21 |
| Appendices | | | | | |
| Informed consent materials | 32 | Model consent form and other related documentation given to participants and authorised surrogates | | | Included in the IRB review |
| Biological specimens | 33 | Plans for collection, laboratory evaluation, and storage of biological specimens for genetic or molecular analysis in the current trial and for future use in ancillary studies, if applicable | | | N/A |

[a] It is strongly recommended that this checklist be read in conjunction with the SPIRIT 2013 Explanation & Elaboration for important clarification on the items.
[b] Indicates page numbers to be completed by authors during protocol development.

Cite as:
