## Appendix 1 (WHO structured abstract)) for "The Effects of AI-Guided Exercise and a Smart Ring on Arterial Stiffness (GONDOR-AS): protocol for a randomized controlled trial"

| | |
|---|---|
| **Primary Registry and ID** | ClinicalTrials.gov: NCT06644014 |
| **Date of Registration** | October 15, 2024 |
| **Secondary ID(s)** | GONDOR-AS |
| **Source(s) of Support** | Oura Health Oy |
| **Primary Sponsor** | Oura Health Oy |
| **Secondary Sponsor(s)** | Kuopio Research Institute of Exercise Medicine (KuLTu) |
| **Contact for Public Queries** | Pauli Ohukainen (Oura Health Oy) |
| **Contact for Scientific Queries** | Pauli Ohukainen (Oura Health Oy); Heikki Pentikäinen (KuLTu) |
| **Public Title** | Reducing Arterial Stiffness with a Smart Ring and AI-Guided Exercise |
| **Scientific Title** | Reducing Arterial Stiffness with a Smart Ring and AI-Guided Exercise (GONDOR-AS): protocol for a randomized controlled trial |
| **Countries of Recruitment** | Finland |
| **Health Condition(s)** | Cardiovascular health, Arterial stiffness, Low cardiorespiratory fitness |
| **Intervention(s)** | 1. AI-based Coaching: Personalized steady-state exercise guidance via Oura Advisor (LLM-based) . 2. HIIT: Supervised cycling. 3. Control: Oura Ring passive monitoring only. |
| **Inclusion/ Exclusion Criteria** | Inclusion: Ages 30-65; sedentary/low activity (<150min moderate or <75min vigorous/week). Exclusion: CAD, diabetes, uncontrolled hypertension, inability to wear a ring. |
| **Study Type** | Interventional; Randomized, parallel-group, 3-arm, blinded outcome assessment |
| **Date of First Enrollment** | October 16 2024 |
| **Target Sample Size** | 165 |
| **Recruitment Status** | Ongoing |
| **Primary Outcome(s)** | Change in carotid-femoral Pulse Wave Velocity (cfPWV) and Change in VO2max (12 weeks) |
| **Key Secondary Outcome(s)** | Chance in Body weight, change in body fat, change in muscle mass (12 weeks) |
| **Ethics Review** | Regional Medical Research Ethics Committee of Eastern Finland Collaborative Area (Approved) |
| **Completion Date** | April 2026 (Estimated) |
| **Summary Results** | Not yet available (data collection and management ongoing) |
| **IPD Sharing Statement** | De-identified participant-level dataset and code to be made public via repository upon publication (pending specific participant permission) |
